# Glomerular Segmentation, Classification, and Pathomic Feature-based Prediction of Clinical Outcomes in Minimal Change Disease and Focal Segmental Glomerulosclerosis

**DOI:** 10.1101/2025.10.01.25336172

**Authors:** Akhil Ambekar, Maryam Roohian, Qian Liu, Bangchen Wang, Fan Fan, Clarissa Cassol, Kyle Lafata, Lawrence Holzman, Laura Mariani, Jeffrey Hodgin, Jarcy Zee, Andrew Janowczyk, Laura Barisoni

**Affiliations:** Division of AI and Computational Pathology, Department of Pathology, Duke University, Durham, North Carolina, USA; Center of Biostatistics and Health Data Science, Brown University, Providence, Rhode Island, USA; Children’s Hospital of Philadelphia Research Institute, Philadelphia, Pennsylvania, USA; Department of Biomedical Engineering, Emory University and Georgia Institute of Technology, Atlanta, Georgia, USA; Arkana Laboratories, Littel Rock, Arkansas, USA; Department of Radiation Oncology, Duke University, Durham, North Carolina, USA; Department of Radiology, Duke University, Durham, North Carolina, USA; Department of Electrical Engineering, Duke University, Durham, North Carolina, USA; Division of Nephrology and Hypertension, Department of Medicine, University of Pennsylvania, Philadelphia, Pennsylvania, USA; Division of Nephrology, Department of Internal Medicine, University of Michigan, Ann Arbor, Michigan, USA; Department of Pathology, University of Michigan, Ann Arbor, Michigan, USA; Department of Biostatistics, Epidemiology, and Informatics, University of Pennsylvania Perelman School of Medicine, Philadelphia, PA, USA; Division of Precision Oncology, Department of Oncology, University Hospital of Geneva, Geneva, Switzerland; Division of Clinical Pathology, Department of Diagnostics, University Hospital of Geneva, Geneva, Switzerland; Division of Nephrology, Department of Medicine, Duke university, Durham, North Carolina, USA

## Abstract

**Background:** Conventional assessment of Focal Segmental Glomerulosclerosis and Minimal Change Disease focuses on the presence/extent of segmental (SS) and global (GS) glomerulosclerosis. While SS and GS represent ongoing and terminal process, encoded in non-SS/GS glomeruli is prognostic information that can be extracted before structural changes are visually discernable. This study applies computational image analysis to (a) automate the segmentation and classification of glomeruli into GS, SS and non-GS/SS, (b) extract subvisual pathomic characteristics from non-GS/SS glomeruli, and (c) assess their clinical relevance.

**Methods:** Leveraging the NEPTUNE/CureGN Periodic acid Schiff-stained whole slide images, we (i) developed deep learning (DL) models for the segmentation and classification of glomeruli into GS, SS and non-GS/SS; (ii) compared the association with disease progression and proteinuria remission of DL-derived percent of GS and SS vs. human scoring; (iii) extracted pathomic features from non-GS/SS; (iv) assessed their prognostic value using ridge-penalized Cox regression, with pathomic features ranked by Maximum Relevance Minimum Redundancy algorithm; and (v) estimated associations between selected pathomic features and clinical outcomes using Cox proportional hazard models.

**Results:** Agreement between computer-aided and visual scoring was good for %GS (ICC = 0.889) and moderate for %SS (ICC = 0.592). The prognostic performance of Cox models of computer-aided visual scoring approaches was comparable (iAUCs 0.779 vs. 0.776 for disease progression and 0.811 vs. 0.817 for complete proteinuria remission, respectively). For non-GS/SS glomeruli, 3 and 4 pathomic features were selected and demonstrated modest prognostic performance for disease progression (iAUC = 0.684) and proteinuria remission (iAUC = 0.661), respectively. After adjusting for demographics, clinical characteristics, %GS and %SS, 2 pathomic features remained statistically significantly associated with proteinuria remission.

**Conclusion:** Computational pathology allows for automatic quantification of SS/GS glomeruli that is comparable to manual assessment for outcome prediction, and the uncovering of previously under-recognized clinically useful information from non-GS/SS glomeruli.

## INTRODUCTION

In patients with focal segmental glomerulosclerosis (FSGS) and minimal change disease (MCD), visual assessment of the extent and qualitative characteristics of segmental (SS) and global glomerulosclerosis (GS) has diagnostic and prognostic implications^1,2,3,4^. However, visual assessment is limited by subjectivity, reduced reproducibility, and an inability to fully quantify the spectrum of sclerotic and non-sclerotic glomerular characteristics.

The advent of digital pathology—enabling the conversion of glass slides into whole-slide images (WSIs)— alongside the development of advanced computational image analysis techniques, including both foundation and traditional methodologies, has created new opportunities for in-depth tissue interrogation^5^. These computational tools offer precise, reproducible quantification of tissue characteristics^5^. While foundation models^6^ provide broad versatility and adaptability, conventional supervised deep-learning (DL) architectures (e.g., convolutional neural network/UNet) excel on defined tasks and yield explicit, quantifiable outputs—for example, segmenting and classifying renal functional tissue units (FTUs) on WSI (e.g., glomeruli, tubular segments, interstitium, arteries, and peritubular capillaries) and detecting specific cell types such as inflammatory cells^2,7–12^. From segmented FTUs, next-generation pathomic (sub-visual)^13^ features can be extracted and quantified, providing novel information that would not be accessible through conventional light microscopy-based diagnostics^5,13–15^: the human eye is not equipped to capture and quantify the full range of their morphological characteristics. Furthermore, while GS and SS reflect a visually discernable stages of glomerular scarring, encoded in non-GS/SS glomeruli may be subtle information relevant to disease progression

The objective of this study is to use computational image analysis to (a) automate the classification and quantification of glomeruli into GS, SS and non-GS/SS; (b) compare the association with clinical outcome of DL-derived and visual scoring-derived percent of GS and SS glomeruli; (c) characterize non-GS/SS segmented glomeruli using pathomics, and (c) assess their clinical relevance to test the hypothesis that non-GS/SS glomeruli harbor prognostically valuable information beyond what is discernible by the human eye.

## METHODS

### Study Dataset Overview

NEPhrotic Syndrome STudy NEtwork (NEPTUNE)^16^ and Cure GlomeruloNephropathy (CureGN)^17^ participants with available digital kidney biopsies were included. Only Periodic acid Schiff (PAS) WSIs were used for this study. (See Supplemental material).

#### NEPTUNE/CureGN clinical and demographic data

Demographics were collected at study enrollment, while medication history, laboratory and clinical data at study enrollment and at each study visit^16,17^. (See Supplemental material). <u>NEPTUNE/CureGN clinical outcomes:</u> Two clinical outcomes were included: (1) time from biopsy to disease progression, defined as a ≥40% decline in estimated glomerular filtration rate (eGFR) with eGFR <90 mL/min per 1.73m^2^ ^18–20^ or kidney failure (chronic dialysis, transplant, or two consecutive eGFRs <15 mL/min per 1.73m^2^); and (2) time from biopsy to first complete proteinuria remission, defined by urine protein creatinine ratio (UPCR) <0.3 g/g. Detailed methods for both outcomes have been previously published^21^.

#### Datasets

Each of the experiments illustrated below were based on different datasets. Thus, description of the dataset is summarized within the pertinent section. A detailed flowchart is shown in Figure 1.

**Figure 1:**
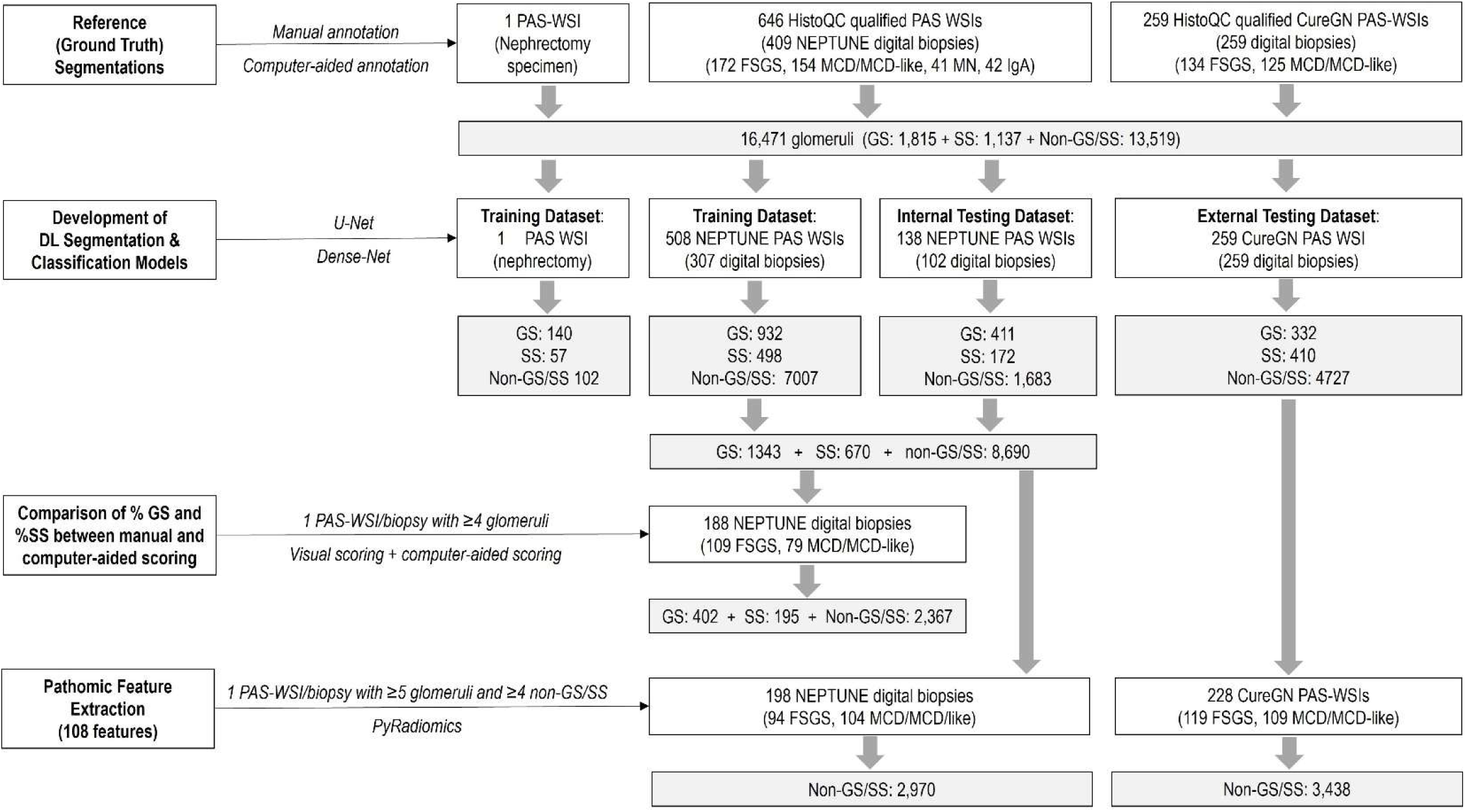
Study overview

### Generation of Reference (Ground Truth) Annotations

#### Dataset

we used (i) a formalin-fixed and paraffin embedded tissue block (Duke pathology archives) from 1 nephrectomy with extensive GS and SS, which was cut at 3 micron and stained with PAS and scanned into a WSI; (ii) 646 PAS-stained WSIs from 409 NEPTUNE biopsies with a diagnosis of MCD (including MCD-like – biopsies without evidence of segmental sclerosis but with percent of GS exceeding that expected for age and/or partial foot process effacement^22^), FSGS, Membranous Nephropathy (MN) and IgA Nephropathy (IgAN); and (iii) 259 PAS-stained WSIs from 259 CureGN biopsies with a diagnosis of FSGS and MCD/MCD-like (Figure 1). For FSGS cases, WSIs with no SS glomerulus were excluded.

#### Reference annotation

Reference annotations were generated using a combination of manual and computer-aided approaches. (See Supplemental Material)

### DL Segmentation and Classification of Glomeruli

#### Dataset

PAS-stained WSIs from the Duke nephrectomy specimen and from the NEPTUNE MCD, FSGS, MN and IgAN digital kidney biopsies were used for training and internal testing of models for the automatic segmentation and classification of glomeruli into GS, SS, and non-GS/SS. CureGN MCD and FSGS PAS WSIs were used as external testing dataset (Figure 1).

#### Training and validation of DL segmentation and classification models

NEPTUNE WSIs were randomly split into 75% training and 25% testing sets (Figure 1). The nephrectomy WSI was also included in the training set. Two U-Net models (GS and SS+ non-GS/SS) were employed (Figure 2) and the segmentation results from both models were subsequently combined. A Dense-Net model was trained to classify the totality of segmented glomeruli into GS, SS, non-GS/SS (Figure 2). The Dense-Net model performance was evaluated on the NEPTUNE testing dataset (internal validation) and on all CureGN cases (external validation).

**Figure 2:**
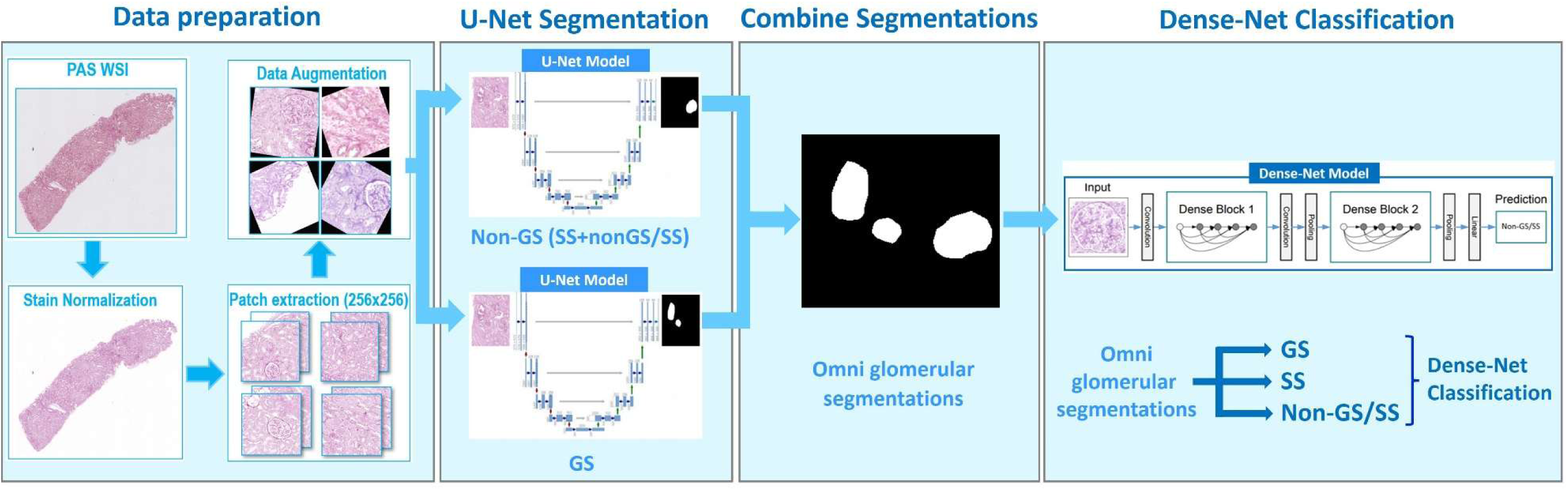
Segmentation and classification of glomeruli: Stain normalization and data augmentation was conducted prior to extraction of patches containing the manually segmented and classified glomeruli. The patches were used to train two U-Net segmentation models, one for GS and SS and one for non-GS/SS, which were then combined into an omni segmentation model. Dense net was used to classify glomeruli into GS, SS, non-GS/SS and non-glomeruli.

#### Evaluation of DL model performance

The evaluation involved computationally counting the true positives (TPs), false positives (FPs), and false negatives (FNs) to calculate overall recall, precision, and F1-score. A glomerulus was considered detected if there was a greater than 50% overlap between the reference annotation and the computationally segmented glomerulus. Additionally, the Dice score, which measures the pixel-level performance of the segmentation model, was computed. Similarly, recall, precision, and F1-score were evaluated for the classification model.

### Comparison of Percent of GS and SS Between the Computer-Aided, Pathologist-QCed Approach and Manual Visual Scoring

#### Dataset

This analysis was restricted to NEPTUNE participants only since NEPTUNE scoring for GS and SS was reported on a 0–100% scale, thus compatible for comparison with computer-aided calculation of percent of GS and SS. In contrast, CureGN core scoring used an ordinal scale and was therefore excluded. NEPTUNE MCD and FSGS participants who had PAS WSIs with four or more complete glomeruli and manual visual scoring at the biopsy level were included. Study participants with less than 4 glomeruli per PAS WSI or missing %GS or %SS values from visual scoring were excluded. (Figure 1).

#### Statistical analysis

Using the computer-aided, pathologist-QCed approach for glomerulus classification on 1 PAS WSI for each study participant, we calculated %GS and %SS as the proportions of total glomeruli on the WSI with GS and SS, respectively. We then compared these percentages to those obtained previously via manual visual scoring at the biopsy level (using multiple WSIs)^23^. Agreement between the two methods was assessed using the intraclass correlation coefficient (ICC).

We then used Cox proportional hazards models to evaluate associations between %GS and %SS and two clinical outcomes: (1) time from biopsy to disease progression, and (2) time from biopsy to first complete proteinuria remission. Models were adjusted for demographic and clinical covariates: age, sex, race, disease cohort, immunosuppressant [IST] use at biopsy, eGFR at biopsy, and UPCR at biopsy. For the disease progression outcome, backward selection was used to reduce the number of covariates to avoid model overfitting due to the limited number of events. Two separate models for each outcome were fitted: one using the computer-aided, pathologist-QCed approach for calculating %GS and %SS on 1 PAS WSI and one using manual visual scoring at the biopsy level. Cox model coefficient estimates and integrated area under the time-varying receiver operating characteristic curve (iAUC)^24^ values derived from each approach were compared between the two approaches.

### Pathomic Feature Extraction from Non-GS/SS Glomeruli

#### Dataset

Participants included in this analysis comprised NEPTUNE and CureGN participants with (1) diagnosis of FSGS or MCD; (2) available clinical outcome data; (3) PAS-stained WSIs that passed HistoQC-driven quality control^25–27^; and (4) at least five glomeruli at the biopsy level and at least four non-GS/SS glomeruli at the WSI level (some biopsies contain more than one WSIs but only one WSI for each participant was used for this analysis). Additionally, for CureGN, we only included participants whose biopsy occurred within three years before enrollment due to missing data between biopsy and study enrollment. We focused pathomic feature analysis on non-GS/SS glomeruli only to test whether encoded in glomerular structures that appear normal to the human eye are information that are clinically relevant.

#### Pathomic feature extraction

A binary mask was utilized to extract the region of interest containing each non-GS/SS glomerulus, excluding non-glomerular regions and the background. Each non-GS/SS glomerulus was represented by a level-8 quantized^28^ glomerular grayscale image. PyRadiomics^28^, a Python package, was employed for feature extraction. A comprehensive set of 108 features, including image intensity (first-order), shape, and texture features, was extracted.

### Prognostic Value of Non-GS/SS-derived Pathomic Features

We tested the association of non-GS/SS pathomic features with 2 clinical outcomes: (1) time from biopsy to disease progression, and (2) time from biopsy to first complete proteinuria remission. Given the limited number of disease progression events, participants from both NEPTUNE and CureGN cohorts were pooled for this analysis. For each participant, glomerulus-level features were aggregated to the patient-level using mean and standard deviation (SD), and patient-level data were used for analysis. To identify the most prognostic pathomic features for each outcome while including demographics and clinical characteristics (age, sex, race, disease cohort, IST use at biopsy, eGFR at biopsy, and UPCR at biopsy), we used the Maximum Relevance Minimum Redundancy (MRMR) algorithm to rank all candidate variables. Ridge-penalized Cox regression was then used to assess prognostic performance as a function of the number of top-ranked variables included. The optimal number of top variables was determined based on the smallest number where the prognostic performance, as measured by iAUC^24^, started to level off.

After identifying the top glomerular pathomic features for each outcome, we evaluated their added prognostic value beyond demographics, clinical characteristics, and computer-generated, pathologist-QCed %GS and %SS values. For each outcome, five models were constructed: (1) a base model including only demographics and clinical characteristics; (2) the base model plus %GS and %SS; (3) a model with only top glomerular pathomic features; (4) top pathomic features added to the base model; and (5) pathomic features added to the base model plus %GS and %SS. Only participants with complete data on all variables used in this analysis were included. All models were fit using ridge regression, and prognostic performance was assessed using iAUC, with internal validation and bias correction via bootstrapping.

Finally, we estimated associations between each selected top pathomic feature and clinical outcome using separate Cox proportional hazard models. For each feature, three models were fitted: (1) unadjusted, (2) adjusted for demographics and clinical characteristics, and (3) further adjusted for computer-generated, pathologist-QCed %GS and %SS.

## RESULTS

### Study dataset

#### Reference (ground truth) glomerular annotations

A total of 16,471 glomeruli from 906 PAS WSIs were segmented (Figure1).

#### WSI dataset for training and testing of the segmentation and classification models

A total of 906 PAS WSIs were used, of which 1 was from the Duke nephrectomy, 646 from 409 NEPTUNE digital biopsies (n= 154 MCD/MCD-like, n= 172 FSGS, n= 41 MN, and n= 42 IgAN), and 259 from 259 CureGN digital biopsies (n= 125 MCD/MCD-like, n= 134 FSGS) (Figure1).

#### Participant dataset for comparing %GS and %SS between the computer-aided, pathologist-QCed approach and manual visual scoring

N=188 NEPTUNE participants were included (n=79 MCD/MCD-Like and n=109 FSGS) (Figure1).

#### Dataset for athomic feature extraction and prediction of outcome

A total of 426 participants were included (104 MCD/MCD-Like and 94 FSGS from NEPTUNE and 109 MCD/MCD-Like and 119 FSGS from CureGN) for a total of 6,408 segmented non-GS/SS glomeruli and corresponding binary masks that were stain normalized and used for feature extraction (Figure1).

Demographics and clinical characteristics of study participants are shown in Table 1, where NEPTUNE participants from two participant datasets described above are combined, and in Supplementary Table 1, where they are presented separately.

**Table 1:** Demographics and clinical characteristics at the time of biopsy, and study outcomes of NEPTUNE and CureGN FSGS/MCD-MCD-like patients.

|  | NEPTUNE<br>(n=233) | CureGN<br>(n=228) |
| --- | --- | --- |
| Age, years | 17.0 (10.0, 42.0) | 22.5 (9.0, 48.0) |
| Children | 121 (52%) | 99 (43%) |
| Adults | 112 (48%) | 129 (57%) |
| Female | 97 (42%) | 119 (52%) |
| Race <sup>a, b</sup> |  |  |
| Black | 62 (28%) | 55 (25%) |
| Other <sup>\$</sup> | 38 (17%) | 25 (11%) |
| White | 125 (56%) | 139 (63%) |
| Hispanic ethnicity <sup>a</sup> | 54 (24%) | 28 (12%) |
| Disease diagnosis |  |  |
| MCD and MCD-Like | 110 (47%) | 109 (48%) |
| FSGS | 123 (53%) | 119 (52%) |
| eGFR <sup>a</sup> | 88.3 (59.5, 109.3) | 91.3 (63.8, 115.6) |
| UPCR <sup>a, b</sup> | 3.1 (1.0, 7.9) | 3.4 (1.0, 8.5) |
| On immunosuppressive medication within 30 days before biopsy or at biopsy | 75 (32%) | 87 (38%) |
| % Global sclerosis <sup>#</sup> | 0.0 (0.0, 12.5) | 0.0 (0.0, 6.1) |
| % Segmental sclerosis <sup>#</sup> | 0.0 (0.0, 8.3) | 0.0 (0.0, 12.5) |
| Follow-up time, years | 3.9 (2.1, 4.6) | 6.5 (4.5, 8.1) |
| Rate of disease progression ($\geq 40\%$ decline in eGFR with eGFR$<90$ or kidney failure) during study follow-up (# of events per 100 person-year) <sup>c</sup> | 5.39 | 4.64 |
| Rate of complete proteinuria remission (UPCR$<0.3$ mg/mg) during study follow-up (# of events per 100 person-year) <sup>d</sup> | 42.49 | 41.48 |

Note: patients included here are NEPTUNE participants (n=233) included in either analysis with glomerular features from non-GS/SS glomeruli (n=198) or analysis on comparing visually scored and computer-aided GS and SS (n=188), and CureGN participants included in analysis with glomerular features from non-GS/SS glomeruli (n=228).
Data are shown as median (IQR), or %(n).
<sup>a</sup> Missing 1% to 5% in NEPTUNE; <sup>b</sup> Missing 1% to 5% in CureGN;
<sup>c</sup> Among n=221 in NEPTUNE and n=227 in CureGN; <sup>d</sup> Among n=189 in NEPTUNE and n=94 in CureGN
<sup>#</sup> Percent of global and segmental sclerosis were based on computer-aided pathologist-QCed approach on single level WSIs.
<sup>\$</sup> Other category includes multi-racial, American Indian /Alaskan Native/First Nation, Asian/Asian American, and Native Hawaiian/Other Pacific Island
IQR: interquartile range; MCD, minimal change disease; FSGS, focal segmental glomerulosclerosis; eGFR, estimated glomerular filtration rate; UPCR, urine protein creatinine ratio

### Performance of DL-based automated segmentation and classification of glomeruli

#### U-Net Detection and Segmentation

There was a high level of accurate detection (recall 92.89% and 93.86%) and precision (92.41% and 93.26%), indicating low FP, and high agreement between the segmented glomeruli and the reference annotations (Dice score 86.42 and 88.97) reflecting the quality of the segmentation results.

Additionally, the F1 score, which considers both precision and recall, was 92.65% (NEPTUNE) and 93.56 (CureGN) (Table 2).

**Table 2:** U-Net Detection and Segmentation of GS, SS and Non-GS/SS Glomeruli for Neptune internal test set (top) and CureGN external test set (bottom)

| Datasets | % Recall | % Precision | % F1-Score | % Dice Score |
| --- | --- | --- | --- | --- |
| Neptune – Internal Test Set | 92.89 | 92.41 | 92.65 | 86.42 |
| CureGN – External Test Set | 93.86 | 93.26 | 93.56 | 88.97 |

#### Dense-Net Glomerular Classification

When analyzing the NEPTUNE internal and CureGN external test dataset, we obtained comparable results: overall F1 scores for GS (91% and 86%) and non-GS/SS (97% for both datasets) were higher compared to SS (76% and 65%). Notably, the F1 score for non-glom class (82%) indicates the model’s ability to differentiate FP predictions that originated from the previous U-Net segmentation results. (Table 3).

**Table 3:** Dense-Net Glomerular Classification – NEPTUNE (top) and CureGN (bottom) by SS, GS, non-GS/SS, and non-glom.

| Class | % Recall | % Precision | % F1-Score | Support (i.e., number of glomerulus or non-gloms) |
| --- | --- | --- | --- | --- |
| NEPTUNE |  |  |  |  |
| SS | 81 | 71 | 76 | 172 |
| GS | 89 | 93 | 91 | 411 |
| non-GS/SS | 96 | 98 | 97 | 1683 |
| non-glom | 89 | 76 | 82 | 205 |
| CureGN |  |  |  |  |
| SS | 66 | 65 | 65 | 410 |
| GS | 86 | 85 | 86 | 332 |
| non-GS/SS | 96 | 97 | 97 | 4727 |
| non-glom | 80 | 71 | 75 | 263 |

### Comparison of Percent of GS and SS Between the Computer-Aided, Pathologist-QCed Approach and Manual Visual Scoring

Among NEPTUNE participants included in this analysis (N=188), the computer-aided approach had good agreement for %GS (ICC = 0.889) with visual scoring and moderate agreement for %SS (ICC = 0.592).

Effect estimates of %GS in Cox proportional hazard models were overall similar (Table 4). For every 10% increase in GS, the hazard of disease progression increased by 1.34 times (95% CI=1.14 to 1.57) based on visual scoring and 1.52 times (95% CI=1.27 to 1.82) based on the computer-aided approach (Table 4a). For complete proteinuria remission, per 10% increase in GS, the hazard of complete remission decreased by 25% (95% CI = 12% to 36%) based on visual scoring and by 23% (95% CI=10% to 34%) based on the computer-aided approach (Table 4b). Effect estimates of %SS were more variable as compared with those from %GS between these two scoring approaches, but still in the same direction and with comparable magnitude.

**Table 4:** Associations between %GS and %SS and clinical outcomes from Cox proportional hazards models, adjusting for demographics and clinical characteristics.

a) Disease progression (N=165)
| Parameter | Visual scoring |  | Computer-aided pathologist-QCed |  |
| --- | --- | --- | --- | --- |
|  | HR (95% CI) | P-value | HR (95% CI) | P-value |
| % GS (per 10 percent) | 1.34 (1.14, 1.57) | 0.0005 | 1.52 (1.27, 1.82) | <.0001 |
| % SS (per 10 percent) | 1.15 (0.90, 1.47) | 0.2582 | 1.39 (1.14, 1.69) | 0.0009 |
| FSGS vs. MCD | 4.48 (1.14, 17.61) | 0.0317 | 2.93 (0.73, 11.80) | 0.1294 |
| UPCR (log 2 transformed) | 1.43 (1.09, 1.89) | 0.0100 | 1.48 (1.10, 1.97) | 0.0090 |
| eGFR at biopsy, per 10 | 0.95 (0.81, 1.12) | 0.5598 | 0.96 (0.82, 1.12) | 0.6147 |
| eGFR at biopsy > 140 | 4.46 (0.80, 24.82) | 0.0879 | 5.38 (0.97, 29.85) | 0.0544 |

\*eGFR winsorized at 140

b) Complete proteinuria remission (N=143)
| Parameter | Visual scoring |  | Computer-aided pathologist-QCed |  |
| --- | --- | --- | --- | --- |
|  | HR (95% CI) | P-value | HR (95% CI) | P-value |
| % GS (per 10 percent) | 0.75 (0.64, 0.88) | 0.0005 | 0.77 (0.66, 0.90) | 0.0007 |
| % SS (per 10 percent) | 0.78 (0.60, 1.03) | 0.0760 | 0.85 (0.71, 1.01) | 0.0617 |
| FSGS vs. MCD | 0.50 (0.28, 0.89) | 0.0183 | 0.45 (0.26, 0.78) | 0.0049 |
| Age at biopsy, years |  |  |  |  |
| ≤ 9 | 0.97 (0.45, 2.09) | 0.9368 | 1.04 (0.48, 2.26) | 0.9194 |
| 10 to 18 | 1.27 (0.57, 2.83) | 0.5600 | 1.33 (0.59, 3.02) | 0.4886 |
| 19 to 43 | 0.70 (0.35, 1.38) | 0.3025 | 0.66 (0.33, 1.32) | 0.2411 |
| 44 to 80 (reference group) |  |  |  |  |
| Female vs. male | 1.14 (0.73, 1.77) | 0.5681 | 1.08 (0.69, 1.68) | 0.7382 |
| Black race | 0.65 (0.34, 1.22) | 0.1782 | 0.70 (0.37, 1.30) | 0.2554 |
| eGFR at biopsy, per 10 units | 0.89 (0.82, 0.98) | 0.0146 | 0.91 (0.83, 0.99) | 0.0369 |
| eGFR at biopsy > 140 | 1.39 (0.54, 3.55) | 0.4934 | 1.42 (0.55, 3.66) | 0.4657 |
| UPCR (log 2 transformed) | 0.96 (0.84, 1.10) | 0.5527 | 0.96 (0.84, 1.10) | 0.5520 |
| IST at biopsy | 1.13 (0.59, 2.14) | 0.7162 | 1.04 (0.55, 1.95) | 0.9155 |

\*eGFR winsorized at 140

The prognostic performance of Cox models using the computer-aided approach was comparable to visual scoring, with iAUCs of 0.779 vs. 0.776 for disease progression and 0.811 vs. 0.817 for complete proteinuria remission, respectively.

### Clinical relevance of pathomic features extracted from non-GS/SS glomeruli

#### Dataset

Pathomic features extracted from non-GS/SS were analyzed to test the association with disease progression in 405 (187 NEPTUNE and 218 CureGN) and with proteinuria remission in 254 (160 NEPTUNE and 94 CureGN) participants, among 426 participants with pathomic features extracted (Figure 1).

#### Model prediction performance

For both outcomes, model prediction performance, as measured by iAUCs, generally increased as the number of MRMR-ranked variables included in the ridge regression model increased. Based on where iAUCs levelled off, six variables (including one shape and two texture glomerular pathomic features) were chosen for the disease progression outcome, including disease cohort (FSGS vs. MCD/MCD-Like), Black race, eGFR at biopsy, SD of perimeter to surface ratio (shape feature), Low Gray Level Run Emphasis (texture features), and Large Area Low Gray Level Emphasis (texture feature). For complete proteinuria remission, nine variables (including four glomerular pathomic features) were chosen: disease cohort (FSGS vs. MCD/MCD-Like), Black race, IST use at biopsy, age at biopsy, mean of informational measure of correlation, eGFR at biopsy, standard deviation of difference average, SD of major axis length, and mean of sphericity.

The top glomerular features alone demonstrated modest prognostic performance for both disease progression (iAUC = 0.684) and proteinuria remission (iAUC = 0.661). When these glomerular features were added to base models including demographics and clinical characteristics, the iAUCs improved slightly from 0.773 to 0.785 for disease progression and from 0.737 to 0.757 for proteinuria remission. When top glomerular features were added to models that also included %GS and %SS, iAUC was similar for disease progression (from 0.798 to 0.802) and increased slightly for proteinuria remission (from 0.757 to 0.769).

#### Association with clinical outcomes

In standard Cox regression models, all three top features had statistically significant associations with disease progression in unadjusted models (Table 5a). However, after adjusting for demographics and clinical characteristics only, or additionally adjusting for %GS and %SS, none remained significant. For proteinuria remission, all four top features had statistically significant associations in unadjusted models. After adjusting for demographics, clinical characteristic, %GS and %SS, two glomerular features remained significant (Table 5b). Every 0.1 unit increase in SD of difference average was associated with 1.18 (95% CI: 1.02-1.35) times higher adjusted hazard of complete proteinuria remission. Every 10 unit increase in SD of major axis length was associated with 12% lower (95% CI: 0.02%-22%) adjusted hazard of complete proteinuria remission. Example biopsies with large and small SD in each of these two glomerular features were shown in Figure 3.

**Figure 3:**
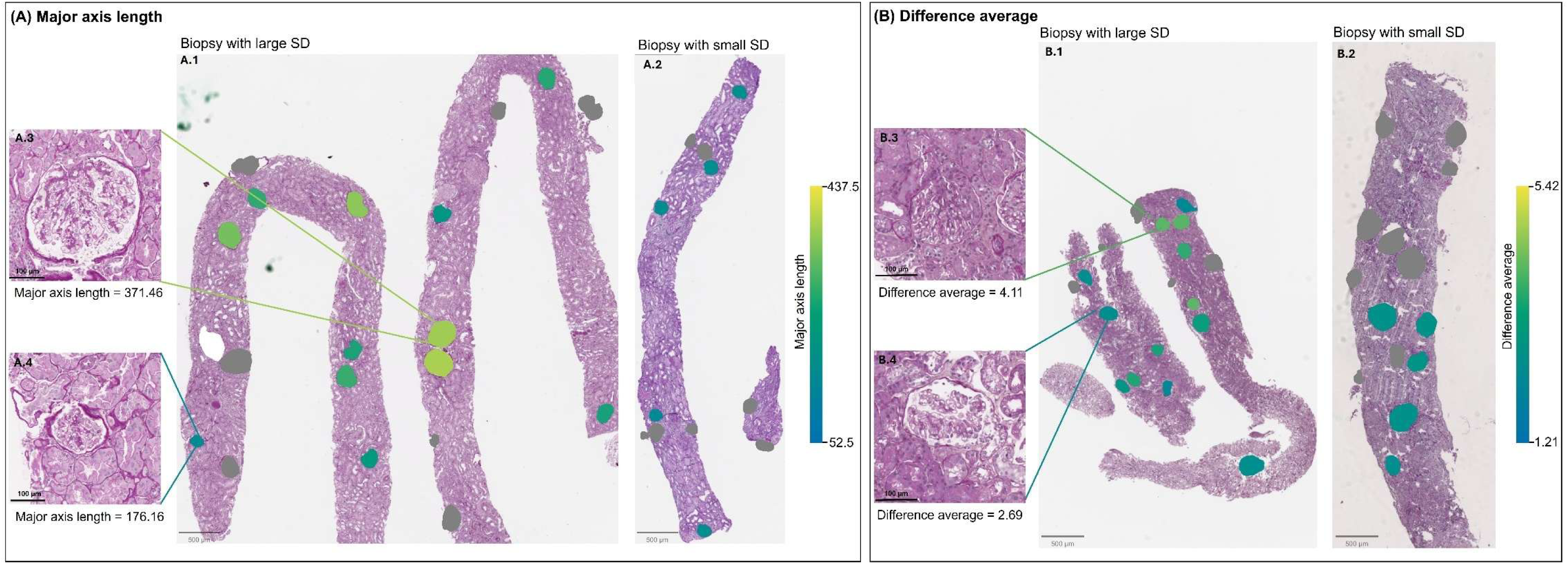
Example biopsies illustrating large and small values of two glomerular features associated with clinical outcome: (A) standard deviation (SD) of major axis length and (B) SD of difference average. In (A), panels A.1 and A.2 represent biopsies with relatively large and small SDs in major axis length, respectively. While panels A.3 and A.4 show individual glomeruli with relatively large and small major axis lengths. In (B), panels B.1 and B.2 represent biopsies with relatively large and small SDs in difference average, respectively, while B.3 and B.4 show individual glomeruli with relatively large and small difference average. A.1 and A.2 are from participants of the same sex, similar age, and similar body mass index (BMI) (A.1: female in mid-20’s, BMI=27 kg/m²; A.2: female in late-20’s, BMI=26 kg/m²). B.1 and B.2 are from participants of the same sex, similar age, and similar BMI (B.1: male in mid-50’s, BMI=25 kg/m²; B.2: male in early-40’s, BMI=23 kg/m²). Biopsies A.1, A.2, B.1. and B.2 are shown at the same magnification. Glomeruli A.3 and A.4 are at the same magnification. Glomeruli B.3 and B.4 are at the same magnification. Glomeruli excluded from analysis (GS, SS, partial, or on the edge glomeruli) are masked in grey.

**Table 5:** Associations between top glomerular features and clinical outcomes from Cox proportional hazards models. Note that all glomerular features reflected glomerulus-level features that were aggregated to the patient-level using summary statistics. Therefore the “mean,” or “standard deviation,” in the feature name refers to the mean and standard deviation across all non-GS/SS glomeruli within a patient, respectively. Demographics and clinical characteristics included age, sex, black race, FSGS vs. MCD, eGFR at biopsy, UPCR at biopsy, and immunosuppressant use at biopsy.

|  | Unadjusted |  |  | Adjusted for demographics and clinical characteristics |  |  | Adjusted for demographics, clinical characteristics, and %GS and %SS |  |  |
| --- | --- | --- | --- | --- | --- | --- | --- | --- | --- |
|  | HR | 95% CI | p-value | HR | 95% CI | p-value | HR | 95% CI | p-value |
| Standard Deviation of Perimeter to Surface Ratio (per 0.001) | 0.87 | (0.80, 0.94) | 0.0009 | 0.92 | (0.84, 1.01) | 0.0965 | 0.93 | (0.85, 1.01) | 0.0909 |
| Mean of Low Gray Level Run Emphasis (per 0.001) | 0.51 | (0.35, 0.75) | 0.0006 | 0.85 | (0.56, 1.27) | 0.4232 | 0.76 | (0.50, 1.16) | 0.1973 |
| Mean of Large Area Low Gray Level Emphasis (per 1) | 1.17 | (1.06, 1.29) | 0.0024 | 1.09 | (0.96, 1.23) | 0.1879 | 1.04 | (0.91, 1.19) | 0.5972 |

b) Complete proteinuria remission
|  | Unadjusted |  |  | Adjusted for demographics and clinical characteristics |  |  | Adjusted for demographics, clinical characteristics, and %GS and %SS |  |  |
| --- | --- | --- | --- | --- | --- | --- | --- | --- | --- |
|  | HR | 95% CI | p-value | HR | 95% CI | p-value | HR | 95% CI | p-value |
| Mean of Informational Measure of Correlation (per 0.01) | 0.93 | (0.90, 0.97) | 0.0005 | 0.96 | (0.93, 1.00) | 0.0848 | 0.97 | (0.93, 1.01) | 0.1751 |
| Standard Deviation of Difference Average (per 0.1) | 1.15 | (1.02, 1.29) | 0.0209 | 1.11 | (0.98, 1.27) | 0.1001 | 1.18 | (1.02, 1.35) | 0.0210 |
| Standard Deviation of Major Axis Length (per 10) | 0.81 | (0.73, 0.90) | 0.0001 | 0.87 | (0.77, 0.98) | 0.0206 | 0.88 | (0.78, 1.00) | 0.0498 |
| Mean of Sphericity (per 0.01) | 1.08 | (1.01, 1.15) | 0.0264 | 1.03 | (0.96, 1.11) | 0.3620 | 1.04 | (0.97, 1.12) | 0.2678 |

## DISCUSSION

This study illustrates a framework for the automated segmentation, classification, and characterization via pathomic analysis of glomeruli in PAS WSIs with the goal of enhancing diagnostic precision and improving prediction of clinical outcomes in glomerular diseases. By leveraging the NEPTUNE and CureGN infrastructure, digital pathology, sophisticated image analysis methodologies, and rigorous pathologist-guided quality control, we developed a pipeline that not only achieved high accuracy in glomerular segmentation and classification of GS, SS, and non-GS/SS, but also identified novel, clinically relevant pathomic features from visually normal-appearing glomeruli (non-GS/SS).

### DL model performance

Our DL segmentation and classification pipeline reached high overall performance across both internal (NEPTUNE) and external (CureGN) datasets. The high Dice and F1-scores for glomerular segmentation reflect excellent agreement with pathologist-generated reference annotations. The lower classification performance for SS compared to GS and non-GS/SS reflects the higher histologic heterogeneity of SS glomeruli and their underrepresentation in the training dataset. These data confirm our previous observations indicating that the number of training objects needed to reach high performance of a DL segmentation model increases proportionately to the heterogeneity in image presentation of the object^2^

### Clinical relevance of manual versus computer-aided scoring

There was good agreement between the traditional manual scoring using all biopsy levels and the automated scoring using a single PAS WSI for %GS (ICC=0.889), with only moderate agreement for %SS (ICC=0.592). However, estimates of effect sizes for both clinical outcomes using computer-aided quantification were of comparable magnitude, directionally consistent, and with prognostic performance similar to those derived from manual scoring. These data suggest that human-machine integrated pipelines can serve as a viable, scalable alternative to labor-intensive manual scoring—especially in the setting of large- clinical research and trials.

### Pathomics uncover hidden glomerular characteristics

Pathomics is the study of tissue characteristics and allows for the extraction and quantification of information that cannot be captured by the human eye. The extraction of a wide range of features from segmented non-GS/SS glomeruli allowed for a detailed understanding of glomerular characteristics, encompassing both structural and textural aspects. The innovative approach of this study is in the exploitation of these sub-visual features from glomeruli that appear normal or near to normal to the human eye, and therefore not accounted for in routine clinical practice. This approach was based on the hypothesis that, if GS represents the end point of glomerular damage and SS an ongoing process, encoded in non-GS/SS are information that may predict the evolution of the damage before it is visible to the human eye.

Glomerular pathomic features for non-GS/SS glomeruli increased prognostic discrimination for complete proteinuria remission even after accounting for demographics, clinical characteristics, and %GS and %SS. Two features—SD of difference average and SD of major axis length—also retained independent associations with remission after adjustment. SD of major axis length indicates variability in glomerular size. Notably, glomerular number, size and size distribution correlate with low birth weight and risk factors for kidney disease^29–31^. Abnormal glomerular size has been recognized in FSGS, with glomerulomegaly commonly observed in post-adaptive forms as a possible mechanism leading to podocyte detachment and segmental scarring^32^, while in other secondary forms, such as mitochondriopathy-associated, the glomerular size is reduced compared to primary or secondary FSGS^33^. Previous studies have shown correlation between glomerular characteristics with serum creatinine (variance) and sex (inverse different moment and textural contrast)^34^. However, there is limited evidence on the direct impacts of size variation on proteinuria remission or other clinical outcomes. These findings highlight the need for additional studies, for example using bulk and spatial omics, to demonstrate the biological plausibility of these observations. Furthermore, SD of difference average reflects variability in textures (i.e., smooth vs. rough/noisy tissue). While this feature may not be directly translatable to known histopathologic descriptors, its significant association with proteinuria remission supports our hypothesis that encoded in non-sclerotic glomeruli are clinically relevant information not detectable through traditional visual assessment. With the rapid advancement of computational methodologies, one can envision future machine-human interactive pipelines deployed in clinical practice enabling a more precise diagnostician and prediction but also the development of new scoring systems with improved visual assessment skill informed by these data-driven discoveries.

### Limitations

Several limitations should be acknowledged, including: (i) the under-representation of SS glomeruli resulting in lower performance of the classifier; (ii) the need of further validation using large international cohorts and other kidney biopsy sources; and (iii) while we demonstrated that there was a good correlation between the computer-aided calculation of %GS and %SS on PAS WSIs versus a more traditional approach using all biopsy levels, further analysis should be performed using a broader representation of tissue sections and stains across each biopsy.

### Summary & Conclusions

This study underscores the value of computational image analysis for reliable and accurate automatic glomerular segmentation, classification, and quantification, and uncovers biomarkers of clinical outcomes from glomeruli lacking overt structural changes. Future work should aim to integrate this data with spatial and non-spatial omics analysis to provide biological context and mechanistic understanding to these observations. These results contribute to the field of glomerular pathology and hold promise for improved risk stratification and personalized treatment strategies in glomerular diseases.

## Supporting information

Glomerular Analysis Manuscript - Supplemental material

## Data Availability

The data in this study were obtained from the Nephrotic Syndrome Study Network (NEPTUNE) and Cure Glomerulonephropathy (CureGN) consortia, where data sharing requires ancillary study approval and a data use agreement. The datasets may be requested from the NEPTUNE Data and Analysis Coordinating Center (DACC), https://neptune-study.org, and the CureGN Data Coordinating Center (DCC), https://www.dev-curegn.org/.

https://www.dev-curegn.org/

## DISCLOSURES

FF and AJ have received financial support from NIH funding list in the acknowledgement. AJ provides consulting for Merck, Lunaphore, and Roche, the latter of which he also has a sponsored research agreement. JZ has received financial support from NIDDK for the submitted work, received grants from NIDDK, NHLBI, NephCure,and Vera Therapeutics, and participates on a Data Safety Monitoring Board for US Renal Care. LM has received financial support from NIDDK and NCATS for the submitted work and received grants from Boehringer-Ingelheim, Travere Therapeutics, Reliant Glycosciences, HiBio, and Takeda Pharmaceuticals in the past 3 years. LM has also received consulting fee from Novartis, Calliditas, Dimerix, Boehringer-Ingelheim, Vera Therapeutics and Travere and payment for educational events from WebMD/Medscape, Otsuka, Apellis and MedLive/PlatformQ. LB has received grants from NIDDK CureGN-Penn PCC, NIDDK Nephrotic Syndrome Rare Disease Clinical Research Network III and NIDDK Computational Pathology for Proteinuric Glomerulopathies, and holds a leadership role in the Scientific Advisory Board of Vertex and NephCure and in the International Society of Glomerular Diseases. LB serves as a consultant for Sangamo, Protalix, Uniquire, and Idorsia. LB has received grants from NIH fundings listed in Acknowledgment, Nephcure and Haller Foundation. LB has also participated on a Data Safety Monitoring Board or Advisory Board for Vertex. JH has received grants from NIH and Department of Defense. CC received honoraria from Novartis as a consultant for the education committee in 2024; is a member of Aurinia advisory board, received funding from SBIR (2023-2024). LH has received grants from NIDDK CureGN-Penn PCC, NIDDK Nephrotic Syndrome Rare Disease Clinical Research Network III, and NIDDK Computational Pathology for Proteinuric Glomerulopathies. Additionally, LH holds a leadership role in the Scientific Advisory Board of NephCure Kidney International.

## ACKNOWLEDGEMENTS

Research reported in this publication was supported by

1. the National Institute of Health (NIH) under the following awards: i) by the National Institute of Diabetes and Digestive and Kidney Diseases (NIDDK) under the award number 2R01DK118431-04; ii) the National Cancer Institute (NCI) under award numbers R01LM013864, R01CA249992-01A1, R01CA202752-01A1, R01CA208236-01A1, R01CA216579-01A1, R01CA220581-01A1, R01CA257612-01A1, 1U01CA239055-01, 1U01CA248226-01, 1U54CA254566-01, and 1U01DK133090; iii) the National Heart, Lung and Blood Institute under award numbers 1R01HL15127701A1, R01HL15807101A1; iv) the National Institute of Biomedical Imaging and Bioengineering under award number 1R43EB028736-01; and v) the National Center for Research Resources under award number 1 C06 RR12463-01,
2. VA Merit Review Award IBX004121A from the United States Department of Veterans Affairs Biomedical Laboratory Research and Development Service.
3. the Office of the Assistant Secretary of Defense for Health Affairs, through i) the Breast Cancer Research Program (W81XWH-19-1-0668), ii) the Prostate Cancer Research Program (W81XWH-15-1-0558, W81XWH- 20-1-0851), iii) the Lung Cancer Research Program (W81XWH-18-1-0440, W81XWH-20-1-0595), iv) the Peer Reviewed Cancer Research Program (W81XWH-18-1-0404, W81XWH-21-1-0345). 4) the Kidney Precision Medicine Project (KPMP) Glue Grant and sponsored research agreements from Bristol Myers-Squibb, Boehringer-Ingelheim, and Astrazeneca.
4. The Nephrotic Syndrome Study Network (NEPTUNE) is part of the Rare Diseases Clinical Research Network (RDCRN), which is funded by the National Institutes of Health (NIH) and led by the National Center for Advancing Translational Sciences (NCATS) through its Division of Rare Diseases Research Innovation (DRDRI). NEPTUNE is funded under grant number U54DK083912 as a collaboration between NCATS and the National Institute of Diabetes and Digestive and Kidney Diseases (NIDDK). Additional funding and/or programmatic support is provided by the University of Michigan, NephCure Kidney International, Alport Syndrome Foundation, and the Halpin Foundation. RDCRN consortia are supported by the RDCRN Data Management and Coordinating Center (DMCC), funded by NCATS and the National Institute of Neurological Disorders and Stroke (NINDS) under U2CTR002818.
5. Additional support was also provided by NephCure and the Henry E. Haller, Jr. Foundation.
6. Funding for the CureGN consortium is provided by U24DK100845, U01DK100846, U01DK100876, U01DK100866, and U01DK100867 from the National Institute of Diabetes and Digestive and Kidney Diseases (NIDDK). Patient recruitment is supported by NephCure. Dates of funding for first phase of CureGN was 9/16/2013-5/31/2019. Dates of funding for the second phase of CureGN was 6/1/2019 - 5/31/2024.
7. This work was supported by the National Science Foundation Graduate Research Fellowship [DGE-2236662 to JR]

## Members of the Nephrotic Syndrome Study Network (NEPTUNE)

### NEPTUNE Collaborating Sites

*Atrium Health Levine Children’s Hospital, Charlotte, SC*: Susan Massengill*, Layla Lo^#^

*Cleveland Clinic, Cleveland, OH*: Katherine Dell*, John O’Toole*, John Sedor**, Victoria Grange^#^

*Children’s Hospital, Denver, CO:* Bradley Dixon*, Nathan Rogers^#^

*Children’s Hospital, Los Angeles, CA*: Rachel Lestz*, Natalie Esquivias^#^

*Children’s Mercy Hospital, Kansas City, MO*: Tarak Srivastava*, Kelsey Markus^#^

*Cohen Children’s Hospital, New Hyde Park, NY*: Christine Sethna*, Suzanne Vento^#^

*Columbia University, New York, NY:* Pietro Canetta*

*Duke University Medical Center, Durham, NC:* Opeyemi Olabisi*, Rasheed Gbadegesin**, Kimberly Cicio^#^

*Emory University, Atlanta, GA:* Laurence Greenbaum*, Chia-shi Wang*, Chris Fan^#^

*The Lundquist Institute, Torrance, CA:* Sharon Adler*, Janine LaPage^#^

*John H Stroger Cook County Hospital, Chicago, IL:* Amatur Amarah*

*Johns Hopkins Medicine, Baltimore, MD:* Meredith Atkinson*, Ryan Hutson^#^

*Mayo Clinic, Rochester, MN:* John Lieske, Marie Hogan, Fernando Fervenza

*Medical University of South Carolina, Charleston, SC:* David Selewski*, Cheryl Alston^#^

*Montefiore Medical Center, Bronx, NY:* Kim Reidy*, Michael Ross*, Frederick Kaskel**, Patricia Flynn^#^

*New York University Medical Center, New York, NY:* Laura Malaga-Dieguez*, Olga Zhdanova**, Laura Jane Pehrson^#^, Melanie Miranda^#^

*The Ohio State University College of Medicine, Columbus, OH*: Salem Almaani*, Laci Roberts^#^

*Riley Children’s Hospital of Indiana University, Indianapolis, IN:* Myda Khalid*, Veronica Servin^#^

*Stanford University, Stanford, CA:* Richard Lafayette*, Elizabeth Chen^#^

*Temple University, Philadelphia, PA:* Iris Lee**

*Texas Children’s Hospital at Baylor College of Medicine, Houston, TX*: Shweta Shah*, Thinh Phan^#^

*University Health Network Toronto:* Heather Reich*, Michelle Hladunewich**, Paul Ling^#^, Martin Romano^#^

*University of California at San Diego, San Diego, CA:* Ambarish Athavale*, Caitlin Carter*, Kristin Zeeb^#^

*University of California at San Francisco, San Francisco, CA*: Paul Brakeman*, Daniel Schrader

*University of Colorado Anschutz Medical Campus, Aurora, CO*: James Dylewski* Nathan Rogers^#^

*University of Kansas Medical Center, Kansas City, KS*: Ellen McCarthy*, Catherine Creed^#^

*University of Miami, Miami, FL:* Alessia Fornoni*, Miguel Bandes^#^

*University of Michigan, Ann Arbor, MI:* Matthias Kretzler*, Laura Mariani*, Zubin Modi*, Amanda Williams^#^, Roxy Ni^#^

*University of Minnesota, Minneapolis, MN:* Patrick Nachman*, Michelle Rheault*, Ariel Langenberger^#^, Brady Wallner^#^

*University of North Carolina, Chapel Hill, NC:* Vimal Derebail*, Keisha Gibson*, Anne Froment^#^, Sharia Warren^#^

*University of Pennsylvania, Philadelphia, PA:* Lawrence Holzman*, Kevin Meyers**, Krishna Kallem^#^, Arielle Swenson^#^

*University of Texas San Antonio, San Antonio, TX*: Samin Sharma**

*University of Texas Southwestern, Dallas, TX:* Elizabeth Roehm*, Kamalanathan Sambandam**, Elizabeth Brown**

*University of Washington, Seattle, WA:* Ashley Jefferson*, Sangeeta Hingorani**, Katherine Tuttle^**§^, Linda Manahan ^#^, Emily Pao^#^, Kelli Kuykendall^§^

*Wake Forest University Baptist Health, Winston-Salem, NC:* Jen Jar Lin**

*Washington University in St. Louis, St. Louis, MO*: Brian Stotter*, Joseph Dumayas^#^

#### Data Analysis and Coordinating Center

*University of Michigan:* Matthias Kretzler*, Brenda Gillespie**, Laura Mariani**, Zubin Modi**, Eloise Salmon**, Howard Trachtman**, Hailey Desmond, Sean Eddy, Damian Fermin, Wenjun Ju, Maria Larkina, Chrysta Lienczewski, Rebecca Scherr, Jonathan Troost, Amanda Williams, Yan Zhai; *Cleveland Clinic:* Crystal Gadegbeku**, John Sedor**, *Duke University:* Laura Barisoni**; *Harvard University:* Matthew G Sampson**; *Northwestern University:* Abigail Smith**; *University of Pennsylvania:* Lawrence Holzman**, Jarcy Zee**

#### Digital Pathology Committee

Carmen Avila-Casado *(University Health Network)*, Serena Bagnasco *(Johns Hopkins University)*, Lihong Bu *(Mayo Clinic)*, Shelley Caltharp *(Emory University)*, Clarissa Cassol *(Arkana)*, Dawit Demeke *(University of Michigan)*, Brenda Gillespie *(University of Michigan)*, Jared Hassler *(Temple University)*, Leal Herlitz *(Cleveland Clinic)*, Stephen Hewitt *(National Cancer Institute)*, Jeff Hodgin *(University of Michigan)*, Danni Holanda *(Arkana)*, Neeraja Kambham *(Stanford University)*, Kevin Lemley, Laura Mariani *(University of Michigan)*, Nidia Messias *(Washington University)*, Alexei Mikhailov *(Wake Forest)*, Vanessa Moreno *(University of North Carolina)*, Behzad Najafian *(University of Washington)*, Matthew Palmer *(University of Pennsylvania)*, Avi Rosenberg *(Johns Hopkins University)*, Virginie Royal *(University of Montreal)*, Miroslav Sekulik *(Columbia University)*, Barry Stokes *(Columbia University)*, David Thomas *(Duke University)*, Ming Wu *(University of New York)*, Michifumi Yamashita *(Cedar Sinai)*, Hong Yin *(Emory University)*, Jarcy Zee *(University of Pennsylvania)*, Yiqin Zuo *(University of Miami)*. Co-Chairs: Laura Barisoni *(Duke University)*, Cynthia Nast *(Cedar Sinai)*.

*Principal Investigator; **Co-investigator; ^#^Study Coordinator; ^§^Providence Medical Research Center, Spokane, WA Last Update: 11SEP2025

##### CureGN Collaborators

The CureGN Consortium members listed below, from within the four Participating Clinical Center networks and Data Coordinating Center, are acknowledged by the authors as Collaborators.

**CureGN PCC Principal Investigators; *CureGN Site Principal Investigators; +CureGN Pathologists, #CureGN Lead Coordinators.

### CureGN Participating Clinical Centers (PCC) through Columbia University

*<u>Columbia University, New York, NY, US:</u>* Gerald Appel, Revekka Babayev, Ibrahim Batal ^+^, Andrew Bomback**, Pietro Canetta, Brenda Chan, Vivette Denise D’Agati ^+^, Samitri Dogra, Hilda Fernandez, Gabriele Gaggero^+^, Ali Gharavi**, William Hines, , Krzysztof Kiryluk**, Satoru Kudose ^+^, Fangming Lin, Victoria Kolupaeva#, Maddalena Marasa, Glen Markowitz ^+^, Mariela Navarro-Torres, Hila Milo Rasouly, Sumit Mohan, Nicola Mongera, Jordan Nestor, Jai Radhakrishnan, Maya Rao, Maya Sabatello, Simone Sanna-Cherchi, Dominick Santoriello^+^, Miroslav Sekulic ^+^, , Michael Barry Stokes^+^, Natalie Uy, Natalie Vena, Benjamin Wooden

*<u>University of Warsaw, Warszawa, Poland:</u>* Bartosz Foroncewicz, Natalia Wiewiórska-Krata, Barbara Moszczuk, Krzysztof Mucha*, Agnieszka Perkowska-Ptasińska, Elżbieta Ryszkowska

*<u>IRCCS Giannina Gaslini, Genoa, Italy:</u>* Francesca Lugani, Valerio Vellone^+^

### CureGN Participating Clinical Centers (PCC) through the Pediatric Nephrology Research Consortium

<u>*Children’s Hospital of New Orleans/ LSU Health, New Orleans, LA, USA:*</u> Diego Aviles*

<u>*Children’s Mercy Hospital, Kansas City, MO, USA:*</u> Tarak Srivastava*, Alexander Katz^+^

<u>*Children’s National Medical Center, Washington DC, USA:*</u> Sun-Young Ahn*

<u>*Cincinnati Children’s Hospital Cincinnati, OH, USA:*</u> Prasad Devarajan, Elif Erkan*, Hillarey Stone

<u>*Connecticut Children’s Medical Center, Hartford, CT, USA:*</u> Sherene Mason*

<u>*East Carolina University Brody School of Medicine, Greenville, NC, USA:*</u> Liliana Gomez-Mendez*

<u>*Emory University, Atlanta, GA, USA:*</u> Larry Greenbaum**, Chia-shi Wang, Hong (Julie) Yin^+^

<u>*Helen DeVos Children’s Hospital, Grand Rapids, MI, USA:*</u> Goebel Jens*

<u>*Levine Children’s Hospital/Atrium Health, Charlotte, NC, USA:*</u> Donald Weaver*

<u>*Lurie Children’s Hospital, Chicago IL, USA:*</u> Jill Krissberg*, Jerome Lane

<u>*Medical College of Wisconsin, Milwaukee, WI, USA:*</u> Cindy Pan, Ellen Cody*

<u>*Nationwide Children’s Hospital, Columbus, OH, USA:*</u> Samantha Martinek-Bundt#, Dawson Carmean#, Mary Dreher^#^, Mahmoud Kallash*, John Mahan**, Samantha Sharpe^#^, William Smoyer**, Laura Biederman^+^

<u>*Oregon Health and Science University, Portland, OR, USA:*</u> Amira Al-Uzri*, Sandra Iragorri

<u>*Riley Children’s Hospital, Indianapolis, IN, USA:*</u> Myda Khalid**

<u>*Cardinal Glennon Children’s Medical Center/ St. Louis University, St. Louis, MO, USA:*</u> Craig Belsha*

<u>*Texas Children’s Hospital, Houston, TX, USA:*</u> Elizabeth Onugha*, Michael Braun, AC Gomez

<u>*Texas Tech Health Sciences Center, Amarillo, TX, USA:*</u> Tetyana Vasylyeva*

<u>*Children’s of Alabama, University of Alabama, Birmingham, AL, USA:*</u> Daniel Feig*

<u>*University of Colorado Children’s Hospital, Colorado, Aurora, CO, USA:*</u> Melisha Hannah*

<u>*University of Kentucky, Lexington, KY, USA:*</u> Aftab Chishti*

<u>*University of Louisville, Louisville, KY, USA:*</u> Jon Klein**

<u>*Holtz Medical Center, University of Miami, Miami, FL, USA:*</u> Chryso Katsoufis, Wacharee Seeherunvong*

<u>*University of Minnesota Children’s Hospital, Minneapolis, MN, USA:*</u> Michelle Rheault**

<u>*University of New Mexico Health Sciences Center, Albuquerque, NM, USA:*</u> Craig Wong*

<u>*University of Oklahoma Health Sciences Center, Oklahoma City, OK, USA:*</u> Qassim Abid*

<u>*University of Virginia, Charlottesville, VA, USA:*</u> John Barcia*, Agnes Swiatecka-Urban

<u>*University of Wisconsin, Madison, WI, USA:*</u> Sharon Bartosh*

<u>*Washington University in St. Louis, St. Louis, MO, USA:*</u> Brian Stotter*, Joseph Gaut ^+^

### CureGN Participating Clinical Centers (PCC) through the University of North Carolina

<u>*Hôpital Maisonneuve-Rosemont, Montreal, Canada:*</u> Louis-Philippe Laurin*, Virginie Royal^+^, Mathieu Latour^+^, Natlie (Natacha) Patey ^+^

<u>*Medical University of South Carolina, Charleston, SC, USA:*</u> Anand Achanti, Milos Budisavljevic*, Vishwajeeth Pasham^+^

<u>*Northwestern University, Chicago, IL, USA:*</u> Cybele Ghossein, Yonatan Peleg*

<u>*Ohio State University, Columbus, OH, USA:*</u> Salem Almaani*, Isabelle Ayoub, Samir Parikh, Brad Rovin, Anjali Satoskar^+^

<u>*University of Chicago, Chicago, IL, USA:*</u> Anthony Chang^+^

<u>*University of Alabama at Birmingham, Birmingham, AL, USA:*</u> Huma Fatima^+^, Jan Novak, Matthew Renfrow, Dana Rizk*

<u>*<u>University of North Carolina Kidney Center, Chapel Hill, NC, USA</u>:*</u> Dhruti Chen, Vimal Derebail**, Ronald Falk**, Keisha Gibson, Dorey Glenn, Susan Hogan, Koyal Jain, J. Charles Jennette^+^, Vanessa Moreno^+^, Amy Mottl, Caroline Poulton^#^, Monica Reynolds, Manish Kanti Saha, Nicole E. Wyatt

<u>*Vanderbilt University, Nashville, TN, USA:*</u> Agnes Fogo^+^, Neil Sanghani*

<u>*Virginia Commonwealth University, Richmond, VA, USA:*</u> Jason Kidd*, Selvaraj Muthusamy^+^

### CureGN Participating Clinical Centers (PCC) through the University of Pennsylvania

*<u>Children’s Hospital of Philadelphia, Philadelphia, PA, USA</u>*: Rebecca Scobell*, Michelle Denburg, Amy Kogon, Kevin Meyers, Madhura Pradhan

*<u>Cleveland Clinic, Cleveland, OH, CA:</u>* Raed Bou Matar*, John O’Toole, John Sedor

*<u>Cohen Children’s Medical Center, New Hyde Park, NY, USA:</u>* Christine Sethna**, Suzanne Vento^#^

*<u>Johns Hopkins University, Baltimore, MD, USA:</u>* Mohamed Atta, Serena Bagnasco^+^, Alicia Neu, John Sperati*

*<u>Lundquist Institute at Harbor-UCLA Medical Center, Torrance, CA, USA:</u>* Sharon Adler*, Tiane Dai, Ram Dukkipati

*<u>Montefiore Medical Center, The Bronx, New York, NY, USA:</u>* Frederick Kaskel, Kaye Brathwaite, Kimberly Reidy*

*<u>New York University, New York, NY, USA:</u>* Laura Malaga-Dieguez*

*<u>Spokane Providence Medical Center, Spokane, WA, USA:</u>* Katherine Tuttle*

*<u>Stanford University, Palo Alto, CA, USA:</u>* Richard Lafayette*, Kamal Fahmeedah, Elizabeth Talley

*<u>Sunnybrook Health Sciences Centre, Toronto, Canada:</u>* Michelle Hladunewich*

*<u>The Hospital for Sick Children, Toronto, Canada:</u>* Rulan Parekh*

*<u>University Health Network, Toronto, Canada:</u>* Carmen Avila-Casado^+^, Daniel Cattran*, Reich Heather, Meherzad Kutky

*<u>University of Miami, Miami, FL, USA:</u>* Yelena Drexler*, Alessia Fornoni

*<u>University of Michigan, Ann Arbor, MI, USA:</u>* Jeffrey Hodgin^+^, Andrea Oliverio*

*<u>University of Pennsylvania, Philadelphia, PA, USA:</u>* Jon Hogan, Lawrence Holzman**, Matthew Palmer ^+^, Gaia Coppock

*<u>University of Pittsburgh School of Medicine, Pittsburgh, PA, USA:</u>* Michael Mortiz, Juhi Kumar*

*<u>University of Washington, Seattle, WA, USA:</u>* Charles Alpers^+^, J. Ashley Jefferson*

*<u>UT Southwestern, Dallas, TX, USA:</u>* Kamal Sambandam, Bethany Roehm*

### Data Coordinating Center (DCC)

*<u>Cedar Sinai Medical Center, Los Angeles, CA, USA</u>*: Cynthia Nast^+^, Jean Hou^+^

*<u>Duke University, Durham, NC, USA</u>*: Laura Barisoni *Cleveland Clinic, Cleveland, OH, USA:* Crystal Gadegbeku**

*<u>Northwestern University, Chicago, IL, USA:</u>* Abigail Smith**

*<u>University of Michigan, Ann Arbor, MI, USA</u>*: Brenda Gillespie, Bruce Robinson, Matthias Kretzler, Zubin Modi, Laura Mariani**

**Steering Committee Chair:** Lisa M. Guay-Woodford, Children’s Hospital of Pennsylvania, Philadelphia, PA, USA

## Notes

### Competing Interest Statement

AJ provides consulting for Merck, Lunaphore, and Roche, the latter of which he also has a sponsored research agreement. JZ participates on a Data Safety Monitoring Board for US Renal Care. LM has received consulting fee from Novartis, Calliditas, Dimerix, Boehringer-Ingelheim, Vera Therapeutics and Travere and payment for educational events from WebMD/Medscape, Otsuka, Apellis and MedLive/PlatformQ. LB holds a leadership role in the Scientific Advisory Board of Vertex and NephCure and in the International Society of Glomerular Diseases. LB serves as a consultant for Sangamo, Protalix, Uniquire, and Idorsia. LB has also participated on a Data Safety Monitoring Board or Advisory Board for Vertex. CC received honoraria from Novartis as a consultant for the education committee in 2024; is a member of Aurinia advisory board, received funding from SBIR (2023-2024). LH holds a leadership role in the Scientific Advisory Board of NephCure Kidney International.

### Author Declarations

The current study was approved by the Duke Health Institutional Review Board (ID: Pro00108417)

