## Supplementary material for "Glomerular Segmentation, Classification, and Pathomic Feature-based Prediction of Clinical Outcomes in Minimal Change Disease and Focal Segmental Glomerulosclerosis": Glomerular Analysis Manuscript - Supplemental material

**Supplemental methods**

Description of NEPTUNE and CureGN consortia: NEPTUNE and CureGN are multi-center, prospective observational cohort studies of patients with a kidney biopsy performed at the time of enrollment (NEPTUNE) and within 5 years before enrollment (CureGN). Additional inclusion criteria for NEPTUNE included having proteinuria >0.5 g/day (or >1.5 g/day after 2014).

NEPTUNE/CureGN clinical and demographic data: Demographics (including race and ethnicity that were self-reported or reported by parents of children) were collected at study enrollment. Medication history, laboratory and other clinical data were collected at study enrollment and at each study visit. Reporting race and ethnicity in both NEPTUNE and CureGN studies was mandated by the US National Institutes of Health, consistent with the Inclusion of Women, Minorities, and Children policy. All NEPTUNE and CureGN participants provided written informed consent (adults) or assent with parental written consent (children). Detailed descriptions of NEPTUNE and CureGN study protocols have been previously published[1,2].

NEPTUNE/CureGN digital pathology repository: Glass slides stained with hematoxylin & eosin, periodic acid Schiff (PAS), trichrome, and silver from kidney biopsies processed at the enrolling centers were centrally scanned (Aperio Scanscope AT2 and Hamamatsu Nano zoomer 2.0 HT) into WSIs at 40X and stored in the NEPTUNE/CureGN digital pathology repository. Only PAS WSIs were used for this study. The PAS WSIs were first curated using open-source software, HistoQC, to identify the presence of batch effects and eliminate poor-quality images with staining and digitization artifacts (such as tissue folding, knife chatter, blurriness, out of focus, bubble, dirt and pen marks)[3,4].

Reference annotation: Reference annotations were generated using a combination of manual and computer-aided approaches. First, using QuPath[5], junior pathologists manually segmented (annotated) and classified GS, SS and non-GS/SS glomeruli on PAS WSIs from the nephrectomy, and 337 NEPTUNE PAS WSIs, which were then reviewed and corrected by a senior pathologist. The segmentation boundaries were defined as follows: for SS and non-GS/SS the boundary was the outer aspect of the Bowman's capsule cutting across the vascular pole or the tubular pole when visible; for GS the boundary was the outer aspect of the Bowman's capsule when present/visible or the outer aspect of the sclerotic glomerulus. Then, a preliminary U-Net segmentation model was trained using the manual annotations and applied to additional HistoQC-qualified WSIs including 309 NEPTUNE and 259 CureGN PAS WSIs. The resulting DL-based glomerular segmentations and their boundaries were manually corrected using QuPath[5] and Quick Annotator (QA)[6] by a senior pathologist. All segmentations were then uploaded to Patch Sorter (PS)[7] for Quality Control (QC) and bulk labeling as GS, SS, or non-GS/SS by a senior pathologist (LB). Only complete glomeruli with at least four capillary loops were included, with partial glomeruli, glomeruli on the float and glomeruli with artifacts excluded.

Training and validation of DL segmentation and classification models: NEPTUNE biopsies with WSIs were randomly split into 75% training and 25% testing sets (Figure 1). The nephrectomy WSI was also included in the training set. To train the DL segmentation network, two U-Net models were employed using all available reference annotations from the training WSI dataset (Figure 2), one for GS glomeruli and the other one for SS + non-GS/SS glomeruli. The segmentation results from both models were subsequently combined. The models were configured with a fixed patch size of 256x256 and an optimal digital magnification of 5X. Training was conducted over 300 epochs, and the hyperparameters (class weights and augmentation parameters) were fine-tuned using a validation set consisting of 20% of the training data. The epoch with the lowest loss on the validation set was selected to generate segmentation results on the NEPTUNE internal testing set. Additionally, a Dense-Net style DL classification network was trained to classify the totality of segmented glomeruli into GS, SS, non-GS/SS, and a class representing false-positive segmentations referred to as "non-glom" (Figure 2). The Dense-Net model utilized the segmented glomeruli as inputs and was trained using a fixed patch size of 256x256 and an optimal digital magnification of 10X. Training of the Dense-Net models was performed over 300 epochs, and the epoch with the lowest loss on the validation set was chosen to generate classification results on the NEPTUNE testing dataset set as internal validation and on all CureGN cases as external validation.

**Supplementary Table 1**: Demographics and clinical characteristics at the time of biopsy, and study outcomes of NEPTUNE and CureGN FSGS/MCD-MCD-like patients. NEPTUNE participants in separate analyses were presented separately.

| | Analysis on comparing visually scored and computer-aided GS and SS (n=188) | Analysis with glomerular features from non-GS/SS glomeruli (n=426) | |
|---|---|---|---|
| | NEPTUNE (N=188) | NEPTUNE (n=198) | CureGN (n=228) |
| Age, years | 19.0 (11.0, 46.5) | 16.0 (8.0, 37.0) | 22.5 (9.0, 48.0) |
| Children | 91 (48%) | 113 (57%) | 99 (43%) |
| Adults | 97 (52%) | 85 (43%) | 129 (57%) |
| Female | 76 (40%) | 84 (42%) | 119 (52%) |
| Race [a, b] | | | |
| Black | 54 (30%) | 53 (28%) | 55 (25%) |
| Other [$] | 29 (16%) | 34 (18%) | 25 (11%) |
| White | 97 (54%) | 104 (54%) | 139 (63%) |
| Hispanic ethnicity [a] | 46 (25%) | 48 (25%) | 28 (12%) |
| Disease diagnosis | | | |
| MCD and MCD-Like | 79 (42%) | 104 (53%) | 109 (48%) |
| FSGS | 109 (58%) | 94 (47%) | 119 (52%) |
| eGFR [a] | 87.3 (56.0, 106.2) | 92.8 (69.2, 112.5) | 91.3 (63.8, 115.6) |
| UPCR [a, b] | 2.8 (1.0, 7.0) | 3.0 (0.9, 8.7) | 3.4 (1.0, 8.5) |
| On immunosuppressive medication within 30 days before biopsy or at biopsy | 55 (29%) | 70 (35%) | 87 (38%) |
| % Global sclerosis [#] | 0.0 (0.0, 16.7) | 0.0 (0.0, 4.8) | 0.0 (0.0, 6.1) |
| % Segmental sclerosis [#] | 0.0 (0.0, 9.5) | 0.0 (0.0, 4.8) | 0.0 (0.0, 12.5) |
| Follow-up time, years | 4.3 (2.6, 4.7) | 4.0 (2.2, 4.6) | 6.5 (4.5, 8.1) |
| Rate of disease progression (≥40% decline in eGFR with eGFR<90 or kidney failure) during study follow-up (# of events per 100 person-year) [c] | 5.67 | 3.59 | 4.64 |
| Rate of complete proteinuria remission (UPCR<0.3 mg/mg) during study follow-up (# of events per 100 person-year) [d] | 37.67 | 50.50 | 41.48 |

Data are shown as median (IQR), or %(n).

[a] Missing 1% to 5% in NEPTUNE; [b] Missing 1% to 5% in CureGN;

[c] Among n=221 in NEPTUNE and n=227 in CureGN; [d] Among n=189 in NEPTUNE and n=94 in CureGN

[#] Percent of global and segmental sclerosis were based on computer-aided pathologist-QCed approach on single level WSIs.

[$] Other category includes multi-racial, American Indian /Alaskan Native/First Nation, Asian/Asian American, and Native Hawaiian/Other Pacific Island

IQR: interquartile range; MCD, minimal change disease; FSGS, focal segmental glomerulosclerosis; eGFR, estimated glomerular filtration rate; UPCR, urine protein creatinine ratio
